# Assessment of presentation patterns of repetitive behaviors in Autism: a cross-sectional video-recording study. Preliminary report

**DOI:** 10.1101/2021.02.20.21252055

**Authors:** Enzo Grossi, Elisa Caminada, Beatrice Vescovo, Tristana Castrignano, Daniele Piscitelli, Giulio Valagussa, Franco Vanzulli

## Abstract

Twenty expert caregivers wearing a body cam recorded 1868 videoclips in 67 autistic subjects along a 3 months close follow-up. A team consisting of a senior child neuro-psychiatrist and a senior psychologist selected 780 of them as expressing repetitive behaviors (RB) and made an empirical classification according to components, complexity, body parts and sensory channels involved, with the aim to understand better the pattern complexity and correlate with autism severity. The RB spectrum for each subject ranged from 1 to 33 different patterns (average= 11.6; S.D.= 6.82). Forty subjects expressed prevalent simple pattern and 27 prevalent complex patterns. No significant differences are found between the two groups according to ADOS score severity. This study represents a first attempt to systematically document expression patterns of RB with a data driven approach. This may provide a better understanding of the pathophysiology, diagnosis, and treatment of RB.

## Introduction

Repetitive behaviors represent one of the pathognomonic expression in autism [Turner, **1999**]. Their variety is particularly large including motor, sensorial, vocal and intellective components. These behaviors are instable over time, are not always present in the same individual and can change in quantity, in quality and in type. The disability correlated to this clinical phenomenology varies along a continuum, from slight to severe and may have a different meaning in different patients. Repetitive use of objects, repetitive activities or ritualism, and repetitive speech are considered as repetitive behaviors.[Cunningham & Schreibman, **2008**; Melo et al., **2019**] Most studies have addressed specifically the motor component of repetitive behavior, i.e. motor stereotypies.

Motor stereotypies denote repetitive, fixed, and purposeless behavioral action that generally stop with distraction. They involve features such as invariance, repetition, as well as a tendency to be inappropriate in nature. [Turner, **1999**]. They are not exclusive to humans, being also observed in many animal species.[Lutz, **2014**].

Although motor stereotypies usually are a sign of severe neuropsychiatric pathologies such as Autism Spectrum Disorder (ASD), serious sensory deafferentation, intellectual disability, and genetic syndrome [Cohen et al., **2005**], they can also be observed in the behavioral repertoire of healthy subjects. We refer to both typically developing children as well as adults who, at times, exhibit, for example, rhythmic swaying of the head and/or the trunk, ritualized behaviors in falling asleep or in frustrating situations. [Langen, Durston, Kas, van Engeland, & Staal, **2011**; Leekam, Prior, & Uljarevic, **2011**]

Primary simple motor stereotypies [Singer, 2009] are reported to occur in roughly 20%-70% of typically developing children, whereas the prevalence of complex motor stereotypies is reported in roughly 3%-4%. [Foster, **1998**; Katherine, **2018**; Lissovoy, **1961**; MacDonald et al., **2007**; Robinson, Woods, Cardona, & Hedderly, **2016**; Sallustro & Atwell, **1978**] However, complex stereotypies in healthy children might be misdiagnosed [Robinson et al., **2016**], and prevalence estimates might be compromised. Notably, even though typically developing children shown motor stereotypies at an earlier age, they tend to diminish over time, i.e., two years old. [Berkson & Tupa, **2000**; MacDonald et al., **2007**]. On the other hand, once stereotypies appear, generally before age three [K. M. Harris, Mahone, & Singer, **2008**], they tend to last for their several years, as evidenced by an extensive longitudinal study carried out on children and adolescents with primary complex motor stereotypies. In this study, 98% of participants continued to maintain the disturbance up to twenty years old. [Oakley, Mahone, Morris-Berry, Kline, & Singer, **2015**]

In children with developmental disabilities, the prevalence of motor stereotypies can be as high as 61%, and even higher (88%) in children with Autism Spectrum Disorder. [Chebli, Martin, & Lanovaz, **2016**] Recently, dddMelo et al. (2019) reported a higher prevalence of motor stereotypies in individuals with ASD and lower intellective quotient (IQ), while gender was not associated with its prevalence.

According to the lexicon of Diagnostic and Statistical Manual of mental disorders fifth edition (DSM-5) [American Psychiatric Association. & American Psychiatric Association. DSM-5 Task Force., **2013**], the broad term repetitive behaviors become “Restricted, repetitive and stereotyped patterns of behavior, interests, and activities” (RRB). They must be present when diagnosing ASD according to the along with the impairments in social-communication/interactions. Particularly, DSM-5 for the RRB domain is polythetic, i.e., two of four RRBs should be present (reviewed in dddBurns and Matson (2017)). RRBs range from stereotyped motor movements to atypical reactions to sensory inputs. [American Psychiatric Association. & American Psychiatric Association. DSM-5 Task Force., **2013**]. Turner (1999) categorized this broad spectrum of behaviors into two classes: (1) ‘‘lower-level’’ characterized by repetition of movement including stereotyped movements, repetitive manipulation of objects and repetitive forms of self-injurious behavior, and (2) ‘‘higher-level’’, which includes object attachments, insistence on sameness, repetitive language, and circumscribed interests. Lower-level behaviors have been found to be associated with lower cognitive abilities, more deficient adaptive skills, and younger chronological age, whereas higher-level behaviors, have been shown to be either of no relationship or positive relationships with the same variables. [Melo et al., **2019**; Turner, **1999**]

Despite their high frequency and strong diagnostic significance within ASD, RRB have not been fully understood due to their broad spectrum of presentation and pattern complexity.[Melo et al., **2019**] Moreover, dddMatson, Dempsey, and Fodstad (2009) suggested that stereotypies could be detected in early life stages as a salient feature of toddlers with autism.

The pathophysiology of RRB is not fully understood (reviewed in dddPeter, Oliphant, and Fernandez (2017)) The disruption of the prefronto-corticobasal ganglia or cortico-striatal thalamo-cortical pathways[Unal, Beverley, Willuhn, & Steiner, **2009**] has been related to stereotypes. The overstimulation of dopaminergic systems has been shown to be associated with the appearance of stereotypies following the intake of levodopa, amphetamine, and cocaine in animal models. [Canales & Graybiel, **2000**; Imeh-Nathaniel et al., **2017**] Motor stereotypies have been elicited in dopamine transporter knock-out mice suggesting the relation between stereotypies and dopamine pathways. [Cinque et al., **2018**] Repetitive behaviors have also been induced, modulating the striatonigral direct circuit in mice models. [Bouchekioua et al., **2018**] Imaging studies in subjects with motor stereotypies found lower levels of GABA, supporting the involvement of the anterior cingulate cortex and the striatum in the pathophysiology of motor stereotypies. [A. D. Harris et al., **2016**] However, studies performed with different functional and structural procedures did not find a consistent pattern of neuroanatomical characteristics [Goldman, O’Brien, Filipek, Rapin, & Herbert, **2013**], highlighting that physiological and functional disruptions should be deeper investigated than anatomical alterations.

From a clinical perspective, several instruments have been proposed to investigate and assess RRB. Recently a panel of experts reviewed twenty-four instruments developed to measure RRB in subjects with ASD. [Scahill et al., **2015**] Several challenges in measuring stereotypies were highlighted, e.g., a variety of clinical presentation in children with ASD, the inter-individual repertoire of stereotypies as well as the prevalence of stereotypies in typically developing children. On the other hand, systematic video observations may overcome questionnaire and interview limitations to investigate stereotypical behaviors. [Goldman et al., **2013**] Specifically, standardized video-recordings can help depict the intricate pattern of RRB commonly observed in ASD. [Goldman et al., **2009**] Recently, dddMelo et al. (2019) suggested describing and characterize stereotypies employing direct observation methods, e.g., video-recording. A better description and understanding of the variety of repetitive behaviors in individuals with ASD would increase the current knowledge of their pathophysiology and the development of better and more appropriate treatment interventions. We hypothesized that using video-recordings in a natural environment during everyday activities would allow classifying RRB systematically. We also hypothesized that ASD severity might be related to the number and quality of RRB displayed. To test these hypotheses, our multiservice Rehabilitative Therapeutic Structure is a good starting point to systematically mapping the entire repertoire of manifested RRB during everyday life activities, with special focus on motor stereotypies. The mapping was performed through systematic videotape analysis. RRB were cataloged into four domains, i.e., motor, sensory, vocal, and intellective as well as according to their structure: simple (involving a single element among one of the domains) or complex (involving more than one elements of the domains).

However, to evaluate and represent all the behaviors classified as repetitive, and not only motor or sensory ones, we expanded and modified the classification by adding behaviors which seemed to correspond to a need for sameness and a series of behaviors that involve complex motoric sequences and verbal production. (MILITERNI et al. 2002). Being aware of the lack of studies analyzing the relationship between the simplicity and complexity of an RB and the severity of autism (ADOS CSS), we planned a protocol aiming to analyze RRB and to determine the correlation between the number and quality of RB to ASD severity through video-recordings, i. e., VICTOR project (VIdeo CaTaloguing Of Repetitive behaviors),

## Materials and Methods

The study involved a group of 67 children and adolescents with a diagnosis of ASD according to the DSM-5 criteria [American Psychiatric Association. & American Psychiatric Association. DSM-5 Task Force., **2013**] and a diagnosis confirmation based on the Autism Diagnostic Observation Scale - 2nd edition criteria (ADOS-2 [Lord et al., **2015**; Randall et al., **2018**]). All participants were residents and/or outpatients of the day center at a Center for the care and rehabilitation of children and adolescents with ASD, multiple deficits, and intellectual disability of different severity. The intellectual disability was classified as mild, moderate, severe, and profound, according to the DSM-5.[American Psychiatric Association. & American Psychiatric Association. DSM-5 Task Force., **2013**]. Trained expert clinicians in developmental psychology performed all clinical and diagnostic evaluations.

Twenty expert healthcare professionals (all female, trained and graduated in special education, with long-standing professional experience at our Institute) wearing a small body-cam (dimension: 88.4 mm x mm x 19.6 mm) placed on the thorax recorded specific stereotypic behavior in an ecological context during everyday life activities of the 67 subjects with ASD over three months with close follow-up. All healthcare professionals were previously trained to minimize their interaction with study participants while recording and to recognize stereotypical patterns.

Each participant was monitored carefully for five consecutive days a week to ensure capturing all the individual RRB. Each time the healthcare professional noticed the onset of stereotypic behavior, she started discretely video-recording unbeknownst to the subject as the session progressed. In a subsequent session, the healthcare professional was asked to evaluate the possibility of interrupting each type of stereotypy through two different extinction interventions: 1) verbal recall and/or proposal of an alternative activity or other appreciated stimulus: 2) physical guidance.[Boyd, McDonough, & Bodfish, **2012**]

Approximately 1800 videos were obtained and later reviewed by an expert educator who selected and assembled them by discarding duplicates, enhancing and standardizing the quality and duration of the recordings as much as possible, thereby obtaining a video for each RRB manifested by the subject. To this end, 780 videos concerning single RRB were selected. The final version of the video library was saved in an appropriate server environment. Fig. 1 shows the diagram flow of the videoclips selection.

**Fig. 1.**
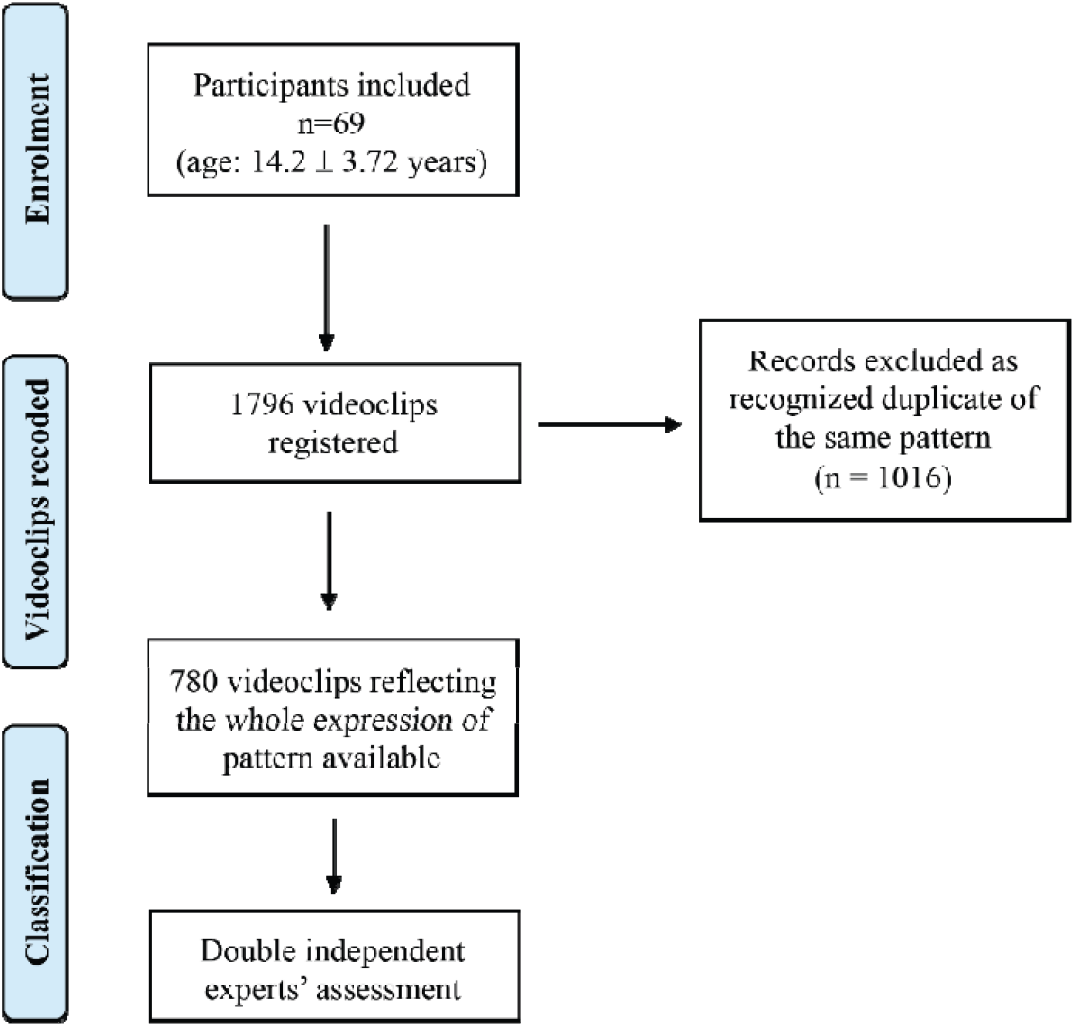
Flow diagram of videoclips selection

The video library was then used as the basis of the study by the team of expert reviewers. The 780 videos (i.e., RRB patterns) obtained were then reviewed and analyzed by an expert team formed by two senior Child Neuropsychiatrists, a senior Developmental Psychologist, and the head of special educators involved in the study. The expert panel described and classified the detected stereotypies. They subsequently compiled the individual grids and loaded them into the database. After detailed discussion and blind agreement on assigning the videos, the behaviors were classified as follows according to four domains, i.e., motor, sensory, vocal and intellective and their quality. Quality was defined as simple (involving a single element among one of the domains) and complex (involving more than one element of the domains). [Militerni, Bravaccio, Falco, Fico, & Palermo, **2002**] The domains were selected based on previous literature. [Goldman et al., **2013**; MacDonald et al., **2007**]

The following codes have been used to classify the components for each RRB:

- Simple Motor (M1): behavior consisting of one movement (e.g., hand flapping or waving, repetitive limb movements, body rocking, etc.);
- Complex Motor (M2): behavior consisting of repeated sequences of movements in several body districts (e.g., toe walking, jumping while walking or running, etc.);
- Simple Sensory (S1): involves a single sensorial aspect (e.g., touching an object or a surface, licking body parts or objects, etc.);
- Complex Sensory (S2): involves different sensory channels (e.g., grasping small items from the floor and put them in the mouth);
- Simple Vocal (V1): repeated simple vocalizations or “noises” (e.g., emitting grunt, raspberries, clearing throat, blowing, etc.);
- Complex Vocal (V2): repeating words phonemes, echolalia, coprolalia;
- Intellective:, rigid, repetitive, stereotyped behaviors that express a need for routine, resistance to change and a tendency to maintain environmental immutability. The Intellective domain was divided between simple and complex behaviors as follows:
- Simple Intellective (I1):. simple rituals of rigid and repetitive behaviors that express a need for routine, resistance to change like for example crumble the food before eating it; always put the glass in the same place, keep the door of the cupboard open in the same way. <u>Complex Intellective</u> (I2): complex ritual from the point of view of the reiterated behavioral sequence. Like for example trashing items while following the same path, line up different object in the same order, etc.

During the coding, the body parts and the sensory component involved were specified. For the body districts considered, please refer to Table 3. The frequency of RRB was reported as the appearance of the behavior one or more times a day. The modality of interruption was classified according to two categories: 1) verbal recall and/or proposal of an alternative activity or other appreciated stimulus. 2) physical guidance by the healthcare professional. Lastly, the prevalent subject state during the appearance of the RRB was defined as quiet, agitated or both.

### Statistical Analysis

Data are presented as number percentages or as means with SDs for nominal and continuous variables, respectively. Pearson correlation (r) analysis was used to investigate correlations as needed; frequency comparison and chi-square tests (χ^2^) were used for categorical variables. Mean comparisons were performed using paired T-tests for paired sample comparisons. Multiple comparisons were adjusted with Bonferroni corrections when appropriate. The level of significance was set at p <0.05.

## Results

Table 1 depicts the demographic characteristics of the participants involved in the study. Thirty-nine (58%) of the subjects had an ADOS-2 calibrated severity score (ADOS-2 CSS) showing high ASD severity. Forty-five subjects (67.2%) had a severe or profound intellectual disability, 19 subjects (28.4%) had moderate intellectual disability, and three subjects (4.4%) had a mild intellectual disability. RRB spectrum ranged from one to 33 different types of patterns (mean= 11.6 ± 6.82; median =10). The most frequent pattern was represented by the combination of simple motor and sensorial components (i.e., M1 S1, accounting for 23% of the total number) followed by a simple motor (M1) and simple sensorial (S1) (9% and 8% respectively). The other 47 patterns with combinations from 1 to 4 components accounted for the remaining 60% with an asymmetric distribution of values. Table 2 shows in detail the frequency and percentage of each stereotypy pattern exhibited during the video-recording.

**Table 1.** Demographic and clinical characteristics of the ASD cohort.

|  |  |
| --- | --- |
| Age range - years | 7.1 - 24.8 |
| Mean Age - years (SD) | 14.2 (3.72) |
| Male N. (%) | 55 (82%) |
| Female N. (%) | 12 (18%) |
| Mean ADOS Total score (SA + RRB) (SD) | 21.14 (5.34) |
| Mean ADOS CSS (SD) | 7.93 (1.72) |
| ADOS CSS - high severity n. subjects (%) | 39 (58%) |
| ADOS CSS - moderate severity n. subjects (%) | 26 (39%) |
| ADOS CSS - low severity n. subjects (%) | 2 (3%) |
| Epilepsy co-occurrence | 10 (15%) |
| Intellectual disability - mild n. subjects (%) | 3 (4.48%) |
| Intellectual disability - moderate n. subjects (%) | 19 (28.36%) |
| Intellectual disability – severe n. subjects (%) | 44 (65.67%) |
| Intellectual disability – profound n. subjects (%) | 1 (1.49%) |

Abbreviations: ADOS CSS, ADOS Calibrated Severity Score

**Table 2.** Distribution of RRB patterns of the sample

| RRB pattern | Frequency | RRB pattern | Frequency |
| --- | --- | --- | --- |
| M1 S1 | 23.08% | **V2** | 0.51% |
| M1 | 8.97% | **M2 S1 I1** | 0.51% |
| S1 | 7.95% | **S2 I1** | 0.38% |
| **M2 S1** | 7.95% | **M2 S2 I1** | 0.38% |
| **I2** | 4.62% | **I2 V1** | 0.38% |
| **M2** | 4.36% | **I1 V2** | 0.26% |
| **M2 S2** | 3.97% | **M2 S2 V2** | 0.26% |
| **S2 M1** | 3.97% | **V2 S1 I1** | 0.26% |
| I1 | 3.59% | **I2 S1 M2** | 0.26% |
| M1 S1 V1 | 3.08% | **M2 V1 I1** | 0.26% |
| **S2** | 2.44% | **S1 V2** | 0.13% |
| **S2 M2 V1** | 2.31% | **M1 V2** | 0.13% |
| I1 M1 | 2.18% | **S2 M1 I1** | 0.13% |
| S1 V1 | 2.18% | **M2 I1 V2** | 0.13% |
| M1 V1 | 1.67% | **M2 V2** | 0.13% |
| **M2 V1** | 1.67% | **M1 S2 V2** | 0.13% |
| **M2 S1 V1** | 1.54% | **I2 S2 V2** | 0.13% |
| S1 I1 | 1.41% | **I2 M2 S1 V1** | 0.13% |
| **S2 M1 V1** | 1.41% | **M2 S1 V2** | 0.13% |
| **I1 M2** | 1.41% | **M2 S2 V2 I1** | 0.13% |
| **S2 V1** | 1.28% | **I2 S2 M2** | 0.13% |
| M1 S1 I1 | 1.15% | I1 M1 V1 | 0.13% |
| **I2 M2** | 0.90% | M1 V1 S1 I1 | 0.13% |
| I2 S1 | 0.90% | **M2 S1 V1 I1** | 0.13% |
| V1 | 0.64% | **M1 V1 I1 S2** | 0.13% |

Abbreviations: Simple motor (M1), complex motor (M2), simple sensorial (S1), complex sensorial (S2), simple Intellective (I1), complex Intellective (I2), simple vocal (V1), complex vocal (V2). In bold complex pattern.

**Table 3.**
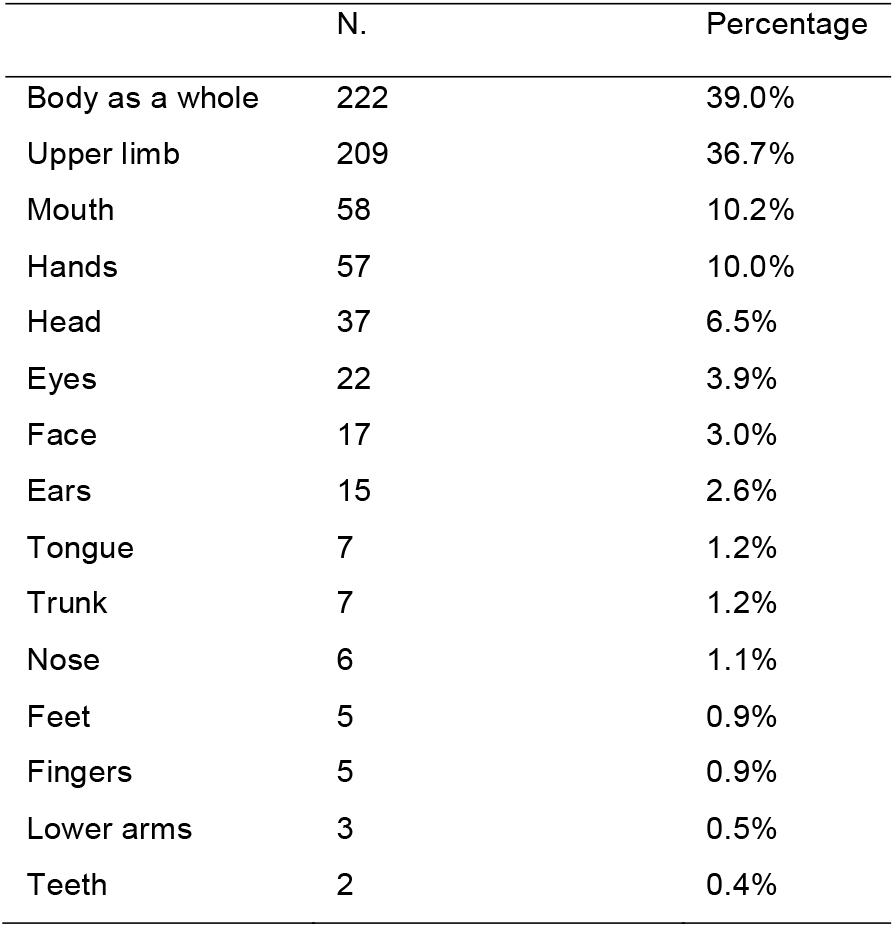
Body parts involved in motor stereotypies

Among the body parts involved during motor stereotypies, the whole body and upper limb accounted for more than 74% (39% and 36.7%, respectively) (Table 3).

Within the 569 out of 780 patterns containing at least one motor component, whole body, and upper limb movements constituted the most common body parts involved (39% and 37% respectively) followed by mouth and hands (10% and 9.8% respectively). Considering the 531 out of 780 patterns containing at least one sensorial component, the most frequent sensory input involved was tactile (50%) followed by proprioceptive (34%) and acoustic (19.5%). Within the 127RRB with vocal components, there were 109 (85.2%) consisting of simple vocalizations and 18 (14.8%) consisting of phonemes or words. The majority (n=681, 87%) of the 780 patterns occurred several times during the day.

Concerning the prevalent state, n=183 (23%) of RRB were shown during both quietness and agitated state, while n=566 (73%) and n= 31 (4%) were exhibited during quietness and agitated state, respectively.

Among the 780 patterns, n= 417 (53.5%), required a physical intervention while n=363 (46.5%) needed a verbal interruption or a proposal of an alternative activity or other stimuli. However, a low and not significant correlation was found between stereotypy pattern and interruption type (verbal or physical). Indeed, maximal correlation values (r) among fifty patterns and interruption modality ranged from -0.08 to +0.08.

Table 4 shows the comparison among subjects with a low number (≤ 5) and a high number (≥ 20) of RRB, not accounting for their complexity features. No significant differences were found between the two groups for age and sex. A significant effect of ADOS-2 CSS (p=0.044) was found on the number of RRB exhibited.

**Table 4.**
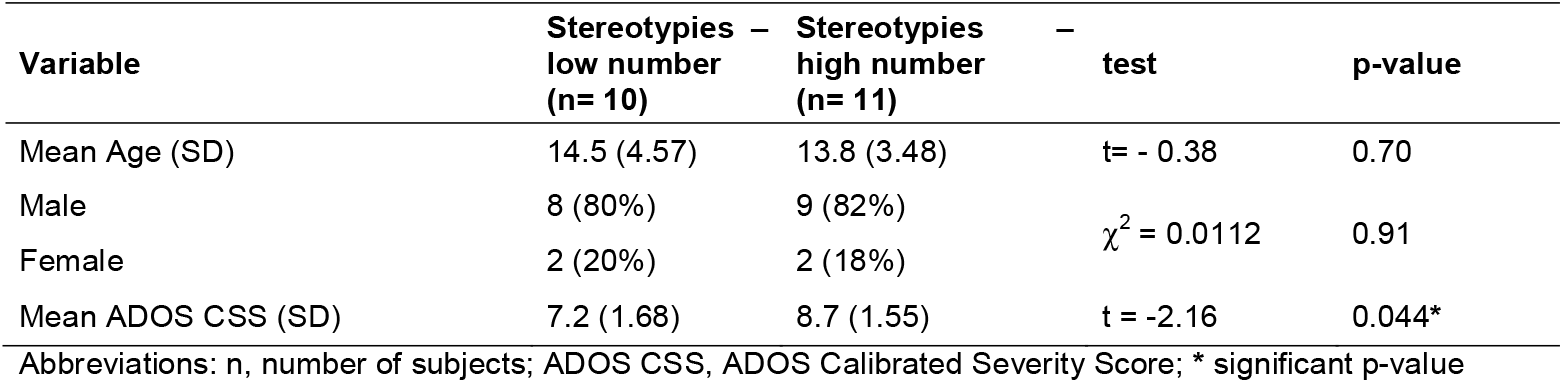
Comparison among subjects with a low number (5 or less) and a high number (20 or more) of stereotypies

To investigate the relation between RRB quality (simple or complex) and autism severity according to the ADOS-2 CSS, a subject was considered to express complex RRB when the proportion of complex over simple patterns exceeded 50%. Therefore, forty subjects (60%) were classified as expressing simple patterns and 27 (40%) participants as expressing complex patterns. Table 5 shows the comparison between subjects with prevalent simple and complex patterns of RRB. No significant differences were found between the two groups respect to age, sex, number of stereotypies, and ADOS-2 CSS.

**Table 5.** Comparison between 40 subjects with a prevalent simple pattern of stereotypies and 27 subjects with a prevalent complex pattern of RRB.

| Variable | Simple pattern group (n= 40) | Complex pattern group (n= 27) | test | p-value |
| --- | --- | --- | --- | --- |
| Mean Age (SD) | 13.87 (3.88) | 13.66 (3.47) | $t = -0.22$ | 0.81 |
| Male | 36 (90%) | 22 (81%) | $\chi^2 = 1.0059$ | 0.32 |
| Female | 4 (10%) | 5 (19%) |  |  |
| Mean number of RRB(SD) | 12.21 (7.82) | 10.85 (5.43) | $t = -0.79$ | 0.43 |
| Mean ADOS CSS (SD) | 7.63 (1.88) | 8.07 (1.26) | $t = 1.06$ | 0.29 |

Abbreviations: ADOS CSS, ADOS Calibrated Severity Score

Table 6 depicts the correlation between the variables examined. Non-significant correlations were found between the total number of RRB who an individual exhibit), age, and ADOS-2 scores for Social Affect subscale. Considering the ADOS-2 CSS as a dependent variable, the number of RRB rather than their complexity features shown the higher correlation values. The ADOS-2 RRB subscale showed a higher correlation coefficient (r=0.458) than ADOS-2 CSS (r=0.347) with respect to the number of subjects with a prevalent complex pattern of RRB.

**Table 6.** Correlation tests among principal variables.

| Variables | No.RRB per subject | No. Simple RRB | No. Complex RRB | Age | ADOS SA | ADOS RRB | ADOS CSS |
| --- | --- | --- | --- | --- | --- | --- | --- |
| No.RRB per subject | 1 |  |  |  |  |  |  |
| No. Simple RRB | **0.809** | 1 |  |  |  |  |  |
| No. Complex RRB | **0.684** | 0.126 | 1 |  |  |  |  |
| Age | 0.102 | 0.0093 | 0.057 | 1 |  |  |  |
| ADOS SA | 0.245 | 0.141 | 0.239 | -0.025 | 1 |  |  |
| ADOS RRB | **0.425** | **0.425** | 0.21 | **0.458** | 0.148 | 1 |  |
| ADOS CSS | **0.376** | **0.376** | 0.232 | **0.347** | 0.094 | **0.757** | 1 |

Abbreviations: N, Number; ADOS SA, ADOS Social Affect subscale; ADOS RRB, ADOS Restricted Repetitive Behavior; ADOS CSS, ADOS Calibrated Severity Score. In bold Pearson (r) values with $p<0.05$ , Bonferroni corrected

## Discussion

Despite their strong diagnostic significance, RRB remain a relatively orphan area in autism research. The lower interest for RRB can be explained mainly to the difficulty in handling the high complexity of existing patterns of presentation.

Rather than classifying RRB into two categories, we have proposed a classification based on a continuum construct according to the complexity of the behavior. Complexity dimensionality is based on the co-occurrence within the single behavior of motor, sensory, vocal and cognitive components, and phenotypic complexity. The latter takes into account the number of body parts or sensory channels involved, the complexity of vocal behavior (sound / phonemes / words) or the sequence of the reiterated ritual.

### Number of RRB and autism severity

We found a significant direct relationship between the number of stereotypes and severity of ASD expressed as ADOS-2 CSS. This result was confirmed by the fact that subjects with a high number of RRB (≥ 20) showed a significantly high ADOS-2 CSS compared to subjects with a low number of RRB (≤ 5). This agrees with several studies that investigated the association between autism severity and RRB. Most of them found that RRB were more frequent as the severity of autism increased. [Akshoomoff, Farid, Courchesne, & Haas, **2007**; Goldman et al., **2009**; Lam & Aman, **2007**; Matson & Rivet, **2008**] On the other hand, dddBodfish, Symons, Parker, and Lewis (2000) reported a non-significant association between RRB and the severity of ASD.

### Pattern of RRB and autism severity

With our careful monitoring of subjects’ behavior, we found that a single subject might exhibit both complex and simple stereotype patterns, suggesting the need to classify at a subject level rather than at the RRB level. Our pragmatic choice was to consider a participant as expressing complex stereotype behavior when the proportion of complex over simple patterns exceeds fifty percent. In our population, 40 subjects were classified accordingly as expressing prevalent simple pattern and 27 as expressing prevalent complex patterns. Interestingly, when we compared the ADOS-2 CSS between subjects with prevalent simple and subjects with prevalent complex patterns of RRB, no significant differences were found. This finding, apparently counterintuitive, might be related to the approach we used for allocating subjects in one of two classes. The poor linear correlation among features of RRB patterns and ADOS-2 CSS should prompt using a machine learning system approach.

Notably, also previous studies that assessed the relationship between RRB and other clinical features in children with ASD found conflicting results. In a large sample of children and adolescents with ASD aged 4–18 years, dddBishop, Richler, and Lord (2006) reported an inverse correlation between ‘‘lower-level’’ RRB and both non-verbal IQ and chronological age, whereas ‘‘higher-level’’ behaviors showed no relationship with IQ. dddMirenda et al. (2010) did not observe any significant relationship between RRB and both non-verbal IQ and chronological age in a sample of 287 preschool-aged children with ASD. dddJoseph, Thurm, Farmer, and Shumway (2013) failed to find a significant relationship between RRB and the non-verbal developmental quotient, chronological age, social communication, and sex in a sample of preschoolers with ASD. In a study of toddlers with ASD, dddWolff et al. (2014) observed that ‘‘higher-level’’ RRB increased with chronological age. They reported that restricted behaviors were modestly negatively correlated with non-verbal developmental quotient at twelve months of age, suggesting that the relationship between RRB and cognitive measures develops over time.

It is worth noting that the heterogeneity of participants with ASD across studies in terms of age, sex, cognitive function, and ASD severity may contribute to obtaining different and occasionally contradictory results and thus may have significantly interfered with a clear understanding of the RRB profile in individuals with ASD. [Melo et al., **2019**] No correlation was found between stereotypy behaviors and interruption interventions, highlighting that the degree of stereotypy complexity was not related to the type of interruption intervention.

### Measuring RRB in individuals with ASD

Previously, dddGoldman and Greene (2012) demonstrate the utility of video-recording to asses repetitive behaviors in preschool children with ASD in a standardized play setting. Recently a panel of expert reviewed questionnaires for assessing repetitive behaviors. [Scahill et al., **2015**] Among the most popular instruments to assess RRB in individuals with ASD there are: The Yale-Brown Obsessive-Compulsive Scale [Goodman, Price, Rasmussen, Mazure, Delgado, et al., **1989**; Goodman, Price, Rasmussen, Mazure, Fleischmann, et al., **1989**], CY-BOCS [Scahill et al., **1997**], the Childhood Routines Inventory [Evans et al., **1997**], the Repetitive Behavior Interview [Turner, **1997**], the Repetitive Behavior Scale-Revised (RBS-R) [Bodfish et al., **2000**], and the ‘‘Restricted interests and repetitive behaviors’’ section of Autism Diagnostic Interview-Revised (ADI-R) [Rutter, LeCouteur, & Lord] All of them are based on caregiver interviews or questionnaires and might suffer from psychometric limitations due to the Likert-based response type. [Piscitelli & Pellicciari, **2018**] To our knowledge, this is the first study that directly investigates RRB and correlates their features with autism severity in an ecological environment. Thanks to this approach, a unique complex scenario appears with the discovery that certain subjects with autism can express more than 30 different patterns of stereotypes.

### Study Limitations and Strengths

This study has some limitations that should be acknowledged. Firstly, results should be interpreted in light of the cross-sectional design of the study and the sample size enrolled. A grouping by age to investigate stereotype longitudinal trajectories was not performed due to the limited sample size that may lead to underpowered subgroup analyses. Video-recordings may be biased due to experimenter-subject interaction. However, health professionals used small bodycams and were trained to interfere as less as possible with the daily clinical routine of the participants. Self-injurious behaviors were not recorded. If a participant showed a harmful behavior (e.g., head banging, self-biting, etc.), the health professionals were trained to stop it and prevent injuries. These limitations were mitigated by the detailed video-recordings carried out during the everyday life of participants.

To this end, this study deserves attention for its strengths: 1) A systematic mapping of the entire repertoire of manifested stereotypes and their video-recording in an ecological context (video analysis); 2) Classification of stereotype according to four behavioral domains (i.e., motor, sensory, vocal and cognitive) and according to their phenotypic complexity, i.e., simple (involving a single motor, sensory, vocal and cognitive domain) or complex (involving multiple domains); 3) An assessment of the relation between stereotyped behavior and sensory disturbances; 4) A proposal to define RRB pattern complexity that could raise a methodological debate on this controversial topic.

## Conclusions

This study contributes to the understanding of repetitive behavior in children and adolescents with ASD. Overall, our findings represent a first attempt to systematically classify the range of repetitive behavior in a cohort of subjects with ASD closely followed by healthcare professionals in an ecological environment. The emerging picture is a detailed description of the broad spectrum of RRB at the individual level. Future research should use a bioinformatics approach such as a machine learning system or applying neural network architectures to classify and follow over time stereotyped behaviors. The scientific community should shape the future research agenda for investigating RRB in health care and in real-world settings. This may provide a better understanding of the pathophysiology, diagnosis, and treatment of RRB. Future research should also investigate the correlation between IQ and RRB. Moreover, studies about older populations with ASD need to be done to elucidate the natural course of RRB in public and private settings.

The data that support the findings of this study are available from the corresponding author (EG) upon reasonable request. Open access to the video bank and to clinical data will be made available to interested researchers, aimed at improving the understanding of this complex phenomenon and its correlation with clinical and demographic features.

## Acknowledgments

The authors thank the special educators who took part in the study performing video-recordings of the stereotypies: Alfiedi Emanuela, Bosisio Gloria, Maspero Veronica, Spreafico Elena, Fratta Antonella, Annoni Valentina, Zanini Elena, Candiani Deborah, Losapio Laura, Colombo Elisabetta, Curioni Elisa, Frassine Alessia, Bernasconi Francesca, Lanzi Giulia, Maspes Serena

## Fundings

This research did not receive any specific grant from funding agencies in the public, commercial, or not-for-profit sectors.

## Conflict of interest

The authors declare that they have no conflict of interest.

## Compliance with Ethical Standards

### Ethical standards

The protocol of this study has been approved by the appropriate ethics committee to which our Institute refers (Comitato Etico dell’Insubria) and therefore the study has been performed in accordance with the ethical standards laid down in the 1964 Declaration of Helsinki and its later amendments.

### Informed Consent

All parents of children and adolescents involved in this study gave their informed consent prior to their inclusion in the study.

## Notes

### Competing Interest Statement

The authors have declared no competing interest.

### Author Declarations

INSUBRIA Ethical Committee gave authorisation.

## References

Akshoomoff, N., Farid, N., Courchesne, E., & Haas, R. (2007). Abnormalities on the neurological examination and EEG in young children with pervasive developmental disorders. J Autism Dev Disord, 37(5), 887–893. doi:10.1007/s10803-006-0216-9

American Psychiatric Association., & American Psychiatric Association. DSM-5 Task Force. (2013). Diagnostic and statistical manual of mental disorders : DSM-5 (5th ed.). Washington, D.C.: American Psychiatric Association.

Berkson, G., & Tupa, M. (2000). Early Development of Stereotyped and Self-Injurious Behaviors. Journal of Early Intervention, 23(1), 1–19. doi:10.1177/10538151000230010401

Bishop, S. L., Richler, J., & Lord, C. (2006). Association between restricted and repetitive behaviors and nonverbal IQ in children with autism spectrum disorders. Child Neuropsychol, 12(4-5), 247–267. doi:10.1080/09297040600630288

Bodfish, J. W., Symons, F. J., Parker, D. E., & Lewis, M. H. (2000). Varieties of repetitive behavior in autism: comparisons to mental retardation. J Autism Dev Disord, 30(3), 237–243. doi:10.1023/a:1005596502855

Bouchekioua, Y., Tsutsui-Kimura, I., Sano, H., Koizumi, M., Tanaka, K. F., Yoshida, K., … Mimura, M. (2018). Striatonigral direct pathway activation is sufficient to induce repetitive behaviors. Neuroscience research, 132, 53–57. doi:10.1016/j.neures.2017.09.007

Boyd, B. A., McDonough, S. G., & Bodfish, J. W. (2012). Evidence-based behavioral interventions for repetitive behaviors in autism. J Autism Dev Disord, 42(6), 1236–1248. doi:10.1007/s10803-011-1284-z

Burns, C. O., & Matson, J. L. (2017). An evaluation of the clinical application of the DSM-5 for the diagnosis of autism spectrum disorder. Expert Rev Neurother, 17(9), 909–917. doi:10.1080/14737175.2017.1351301

Canales, J. J., & Graybiel, A. M. (2000). A measure of striatal function predicts motor stereotypy. Nat Neurosci, 3(4), 377–383. doi:10.1038/73949

Chebli, S. S., Martin, V., & Lanovaz, M. J. (2016). Prevalence of stereotypy in individuals with developmental disabilities: A systematic review. Review Journal of Autism and Developmental Disorders, 3(2), 107-118 %@ 2195-7177.

Cinque, S., Zoratto, F., Poleggi, A., Leo, D., Cerniglia, L., Cimino, S., … Adriani, W. (2018). Behavioral Phenotyping of Dopamine Transporter Knockout Rats: Compulsive Traits, Motor Stereotypies, and Anhedonia. Front Psychiatry, 9, 43. doi:10.3389/fpsyt.2018.00043

Cohen, D., Pichard, N., Tordjman, S., Baumann, C., Burglen, L., Excoffier, E., … Heron, D. (2005). Specific genetic disorders and autism: clinical contribution towards their identification. J Autism Dev Disord, 35(1), 103–116. doi:10.1007/s10803-004-1038-2

Cunningham, A. B., & Schreibman, L. (2008). Stereotypy in Autism: The Importance of Function. Res Autism Spectr Disord, 2(3), 469–479. doi:10.1016/j.rasd.2007.09.006

Evans, D. W., Leckman, J. F., Carter, A., Reznick, J. S., Henshaw, D., King, R. A., & Pauls, D. (1997). Ritual, habit, and perfectionism: the prevalence and development of compulsive-like behavior in normal young children. Child Dev, 68(1), 58-68. Retrieved from http://www.ncbi.nlm.nih.gov/pubmed/9084125

Foster, L. G. (1998). Nervous habits and stereotyped behaviors in preschool children. Journal of the American Academy of Child & Adolescent Psychiatry, 37(7), 711–717. doi:10.1097/00004583-199807000-00011

Goldman, S., & Greene, P. E. (2012). Stereotypies in autism: a video demonstration of their clinical variability. Front Integr Neurosci, 6, 121. doi:10.3389/fnint.2012.00121

Goldman, S., O’Brien, L. M., Filipek, P. A., Rapin, I., & Herbert, M. R. (2013). Motor stereotypies and volumetric brain alterations in children with Autistic Disorder. Res Autism Spectr Disord, 7(1), 82–92. doi:10.1016/j.rasd.2012.07.005

Goldman, S., Wang, C., Salgado, M. W., Greene, P. E., Kim, M., & Rapin, I. (2009). Motor stereotypies in children with autism and other developmental disorders. Dev Med Child Neurol, 51(1), 30–38. doi:10.1111/j.1469-8749.2008.03178.x

Goodman, W. K., Price, L. H., Rasmussen, S. A., Mazure, C., Delgado, P., Heninger, G. R., & Charney, D. S. (1989). The Yale-Brown Obsessive Compulsive Scale. II. Validity. Arch Gen Psychiatry, 46(11), 1012–1016. doi:10.1001/archpsyc.1989.01810110054008

Goodman, W. K., Price, L. H., Rasmussen, S. A., Mazure, C., Fleischmann, R. L., Hill, C. L., … Charney, D. S. (1989). The Yale-Brown Obsessive Compulsive Scale. I. Development, use, and reliability. Arch Gen Psychiatry, 46(11), 1006–1011. doi:10.1001/archpsyc.1989.01810110048007

Harris, A. D., Singer, H. S., Horska, A., Kline, T., Ryan, M., Edden, R. A., & Mahone, E. M. (2016). GABA and Glutamate in Children with Primary Complex Motor Stereotypies: An 1H-MRS Study at 7T. AJNR Am J Neuroradiol, 37(3), 552–557. doi:10.3174/ajnr.A4547

Harris, K. M., Mahone, E. M., & Singer, H. S. (2008). Nonautistic motor stereotypies: clinical features and longitudinal follow-up. Pediatr Neurol, 38(4), 267–272. doi:10.1016/j.pediatrneurol.2007.12.008

Imeh-Nathaniel, A., Rincon, N., Orfanakos, V. B., Brechtel, L., Wormack, L., Richardson, E., … Nathaniel, T. I. (2017). Effects of chronic cocaine, morphine and methamphetamine on the mobility, immobility and stereotyped behaviors in crayfish. Behav Brain Res, 332, 120–125. doi:10.1016/j.bbr.2017.05.069

Joseph, L., Thurm, A., Farmer, C., & Shumway, S. (2013). Repetitive behavior and restricted interests in young children with autism: Comparisons with controls and stability over 2 years. Autism Research, 6(6), 584–595. doi:10.1002/aur.1316

Katherine, M. (2018). Stereotypic movement disorders.

Lam, K. S., & Aman, M. G. (2007). The Repetitive Behavior Scale-Revised: independent validation in individuals with autism spectrum disorders. J Autism Dev Disord, 37(5), 855–866. doi:10.1007/s10803-006-0213-z

Langen, M., Durston, S., Kas, M. J., van Engeland, H., & Staal, W. G. (2011). The neurobiology of repetitive behavior: …and men. Neurosci Biobehav Rev, 35(3), 356–365. doi:10.1016/j.neubiorev.2010.02.005

Leekam, S. R., Prior, M. R., & Uljarevic, M. (2011). Restricted and repetitive behaviors in autism spectrum disorders: a review of research in the last decade. Psychol Bull, 137(4), 562–593. doi:10.1037/a0023341

Lissovoy, V. d. (1961). Head banging in early childhood: A study of incidence. The Journal of Pediatrics, 58(6), 803–805. doi:10.1016/S0022-3476(61)80135-2

Lord, C., Rutter, M., DiLavore, P. C., Risi, S., Gotham, K., Bishop, S. L., & Schedule, A. A. D. O. (2015). ADOS-2. Manual (Part I): Modules, 1–4.

Lutz, C. K. (2014). Stereotypic behavior in nonhuman primates as a model for the human condition. ILAR J, 55(2), 284–296. doi:10.1093/ilar/ilu016

MacDonald, R., Green, G., Mansfield, R., Geckeler, A., Gardenier, N., Anderson, J., Sanchez, J. (2007). Stereotypy in young children with autism and typically developing children. Res Dev Disabil, 28(3), 266–277. doi:10.1016/j.ridd.2006.01.004

Matson, J. L., Dempsey, T., & Fodstad, J. C. (2009). Stereotypies and repetitive/restrictive behaviours in infants with autism and pervasive developmental disorder. Dev Neurorehabil, 12(3), 122–127. doi:10.1080/17518420902936730

Matson, J. L., & Rivet, T. T. (2008). Characteristics of challenging behaviours in adults with autistic disorder, PDD-NOS, and intellectual disability. J Intellect Dev Disabil, 33(4), 323–329. doi:10.1080/13668250802492600

Melo, C., Ruano, L., Jorge, J., Pinto Ribeiro, T., Oliveira, G., Azevedo, L., & Temudo, T. (2019). Prevalence and determinants of motor stereotypies in autism spectrum disorder: A systematic review and meta-analysis. Autism, 1362361319869118. doi:10.1177/1362361319869118

Militerni, R., Bravaccio, C., Falco, C., Fico, C., & Palermo, M. T. (2002). Repetitive behaviors in autistic disorder. Eur Child Adolesc Psychiatry, 11(5), 210–218. doi:10.1007/s00787-002-0279-x

Mirenda, P., Smith, I. M., Vaillancourt, T., Georgiades, S., Duku, E., Szatmari, P., Pathways in, A. S. D. S. T. (2010). Validating the Repetitive Behavior Scale-revised in young children with autism spectrum disorder. J Autism Dev Disord, 40(12), 1521–1530. doi:10.1007/s10803-010-1012-0

Oakley, C., Mahone, E. M., Morris-Berry, C., Kline, T., & Singer, H. S. (2015). Primary complex motor stereotypies in older children and adolescents: clinical features and longitudinal follow-up. Pediatr Neurol, 52(4), 398–403 e391. doi:10.1016/j.pediatrneurol.2014.11.002

Peter, Z., Oliphant, M. E., & Fernandez, T. V. (2017). Motor Stereotypies: A Pathophysiological Review. Front Neurosci, 11, 171. doi:10.3389/fnins.2017.00171

Piscitelli, D., & Pellicciari, L. (2018). Responsiveness: is it time to move beyond ordinal scores and approach interval measurements? Clin Rehabil, 32(10), 1426–1427. doi:10.1177/0269215518794069

Randall, M., Egberts, K. J., Samtani, A., Scholten, R. J., Hooft, L., Livingstone, N., Williams, K. (2018). Diagnostic tests for autism spectrum disorder (ASD) in preschool children. Cochrane Database Syst Rev, 7, CD009044. doi:10.1002/14651858.CD009044.pub2

Robinson, S., Woods, M., Cardona, F., & Hedderly, T. (2016). Intense Imagery Movements (IIM): More to motor stereotypies than meets the eye. Eur J Paediatr Neurol, 20(1), 61–68. doi:10.1016/j.ejpn.2015.10.006

Rutter, M., LeCouteur, A., & Lord, C. Autism Diagnostic Interview-Revised Manual. 2003. Los Angeles: Western Psychological Services.

Sallustro, F., & Atwell, C. W. (1978). Body rocking, head banging, and head rolling in normal children. J Pediatr, 93(4), 704–708. doi:10.1016/s0022-3476(78)80922-6

Scahill, L., Aman, M. G., Lecavalier, L., Halladay, A. K., Bishop, S. L., Bodfish, J. W., Dawson, G. (2015). Measuring repetitive behaviors as a treatment endpoint in youth with autism spectrum disorder. Autism, 19(1), 38–52. doi:10.1177/1362361313510069

Scahill, L., Riddle, M. A., McSwiggin-Hardin, M., Ort, S. I., King, R. A., Goodman, W. K., … Leckman, J. F. (1997). Children’s Yale-Brown Obsessive Compulsive Scale: reliability and validity. J Am Acad Child Adolesc Psychiatry, 36(6), 844–852. doi:10.1097/00004583-199706000-00023

Singer, H. S. (2009). Motor stereotypies. Semin Pediatr Neurol, 16(2), 77–81. doi:10.1016/j.spen.2009.03.008

Turner, M. (1997). Towards an executive dysfunction account of repetitive behaviour in autism. In J. Russell (Ed.), Autism as an executive disorder (pp. 57–100). New York, NY: Oxford University Press.

Turner, M. (1999). Annotation: Repetitive Behaviour in Autism: A Review of Psychological Research. Journal of Child Psychology and Psychiatry, 40(6), 839–849. doi:10.1111/1469-7610.00502

Unal, C. T., Beverley, J. A., Willuhn, I., & Steiner, H. (2009). Long-lasting dysregulation of gene expression in corticostriatal circuits after repeated cocaine treatment in adult rats: effects on zif 268 and homer 1a. Eur J Neurosci, 29(8), 1615–1626. doi:10.1111/j.1460-9568.2009.06691.x

Wolff, J. J., Botteron, K. N., Dager, S. R., Elison, J. T., Estes, A. M., Gu, H., Network, I. (2014). Longitudinal patterns of repetitive behavior in toddlers with autism. J Child Psychol Psychiatry, 55(8), 945–953. doi:10.1111/jcpp.12207

